# Combining centralized and decentralized approaches to assess and ensure data quality in Eurocrine® via Microsoft Power BI and DataquieR

**DOI:** 10.64898/2026.06.04.26354884

**Authors:** TJ Musholt, T Clerici, A Bergenfelz, CO Schmidt, S Struckmann, the Eurocrine Board

## Abstract

**Background:** Medical registries have gained importance in the evaluation of healthcare quality outcomes. In the absence of high-quality evidence, such as randomized controlled trials, studies based on registry data are essential for informing clinical guidelines. Methods for assessing data quality are rarely described in detail. To ensure the credibility of registry-based studies, registries must use all available technical and operational means to guarantee high data quality.

**Method:** Eurocrine^®^ is a pan-European endocrine surgical database and quality registry initially funded by the EU healthcare programme, which started in 2015 and now includes more than 200,000 interventions as of April 2025. To ensure high data quality, interactive and standardized reports are created via Microsoft Power BI, which are created both centrally and locally. In addition, comprehensive data quality analyses were performed via the R-based package dataquieR.

**Results:** Although a multitude of technical measures (for example, input screen design and real-time plausibility checks during data entry) are in place, they are not sufficient to prevent human errors at data entry. Errors identified in the reports were corrected, and preventive measures were implemented. Overall, the data quality was assessed as very good in terms of completeness, accuracy, and consistency.

**Conclusion:** It is very important to provide registry users with an efficient and smart tool to identify data issues, as they have the clinical information to correct them. Data quality reports generated with dataquieR represent an effective tool for registry administrators. Predesigned Microsoft Power BI reports enable participating Eurocrine^®^ clinics to self-audit their data.

## Introduction

Over the past 20 years, an increasing number of medical registries have been established to monitor the quality of care and optimize the treatment of a wide variety of diseases, especially those that cannot be addressed by prospective randomized trials due to low case numbers. A PubMed search using the term “medical registry” revealed a marked increase in the number of related publications, from 753 in 2002 to approximately 14,000 per year between 2020 and 2023. Analyses of medical registry data significantly impact the development of clinical guidelines. For example, the American Thyroid Association (ATA) recommendations for hemithyroidectomy for differentiated thyroid cancer (DTC) < 4 cm are mainly based on analyses of the Surveillance, Epidemiology, and End Results (SEER) registry [1].

The benefit of medical registries largely depends on the quality of the collected data, typically defined by criteria such as completeness, correctness, and relevance [2]. However, both manual and automatic data collection are associated with an error rate of up to 10% to 20%, such as typing errors, mix-up of sides, sex, inconsistent entries, and others [3]. This underlines that measures to assess and ensure data quality are indispensable. The training and motivation of staff involved in data entry is another key factor impacting data quality, which may vary substantially within and between registries or data collection centers.

Guidelines for evaluating the quality of registries have been published, including the EMA’s Guideline on registry-based studies [4], the EUnetHTA’s Registry Evaluation and Quality Standards Tool (REQueST) [5], and the US Agency for Healthcare Research and Quality (AHRQ)’s Users’ Guide on registries [6]. However, statements on data quality and measures undertaken to ensure it are often absent from medical publications [7], including reports of randomized controlled trials [8]. The contributing factors include the lack of generally applicable tools for assessing data quality, along with frequently insufficient financial resources for data collection, and quality assurance, such as active follow-up by trained staff, internal and external auditing, data validation, and appropriate infrastructure maintenance [7]. This illustrates that more guidance is needed to implement efficient and sustainable data quality assurance processes, even when resources are limited.

One method to assess data quality is source data verification, in which electronically recorded data are compared against the original data sources via a two-person, “four-eyes” principle. However, for large registries, this approach is not feasible. A two-step approach seems more feasible: first, suitable tools are used to either search for data anomalies or to randomly select data entries, which can then be verified in a second stage by checking the source data (=re-abstracting). Missing data can be filled in, and false data entries can be corrected. If adequate financial and human resources are available, although this is rarely the case, trained data clerks can also be deployed on site [9]. While the previously mentioned two-step approach provides several options, practical implementation has rarely been addressed, which is particularly challenging for multicenter data collection, where immediate feedback on data anomalies or missing data to individual clinics or users is typically lacking.

This paper therefore describes the implementation of a multi-stage centralized as well as decentralized data quality assurance and assessment pipeline in Eurocrine^®^, a major pan-European endocrine surgery registry. This example may guide other registries to implement extensive data quality assurance activities. In addition, we provide insight into the handling of detected data issues.

## Methods

The setup of the Eurocrine^®^ was initially funded by the EU Health Programme and Region Skane, Sweden. The registry has been fully functional since 2015 and is managed by the EUROCRINE Society, which is legally based in Vienna, Austria, https://eurocrine.eu. The aim of Eurocrine^®^ is to improve the surgical treatment of endocrine tumors and diseases, with a special focus on rare endocrine tumors. A second aim is to enable quality control tools for its member clinics. Since 2022, the registry has played a key role in the accreditation process for European reference centers for endocrine surgery, which is overseen by the European Society or Endocrine Surgeons (ESES).

As of 2025, it includes data on over 200,000 patients, with more than 205,000 endocrine procedures. Thyroid resections (132,000) and operations for hyperparathyroidism (24,000) are the most frequently registered procedures. Additional modules cover procedures for adrenal diseases, paragangliomas and neuroendocrine tumours of the gastrointestinal tract (GEP-NETs). Currently, 161 specialized endocrine surgical units from 22 European countries use the database to document and analyse their patients’ operations and outcomes.

Participation in the registry is voluntary for both individual clinics and national societies representing endocrine surgery in their country. Participating clinics and national societies pay annual fees, which also cover the Microsoft Power BI licence required to analyse and visualize clinic data and aggregated national data. The registry and database are funded through these membership fees.

Data entry is predominantly performed by the treating physician (> 70%) at the time of treatment via a secure web-user interface (GUI) that ensures standardized input. The GUI is available in nine languages to ensure the correct understanding of the data queries. A pseudonymous ID is automatically assigned to each patient during data entry, with the actual identity stored only in the member clinic. Data protection is handled in accordance with the general data protection regulation (GDPR), which must be confirmed for each registration by the user before data entry. Strong two-factor authentication is used for log-in.

Comprehensive measures have been implemented to review and correct the data and to flag incomplete data or data anomalies at various levels. Below, we describe the measures used for the thyroid Eurocrine^®^ module in detail. This module contains a total of 527 variables, of which 40 are mandatory and 11 are automatically derived from specific mandatory variables by filtering and combining categories. The remaining 476 are optional, and most of them capture follow-up data. Each participating country can create additional variables for display on the web interface. Furthermore, participating clinics can define their own variables using an integrated tool named “MyEurocrine”. Not all variables apply to all patients; e.g., variables for benign diseases differ from those for malignant diseases. Therefore, some variables depend on the selected higher-level selections.

All variables are periodically reviewed by the Eurocrine^®^ Data Management Committee (DMC) to ensure the relevance of existing variables, identify potentially missing information, and evaluate overall data quality. The relevance of the data for research purposes is discussed biannually in the joint Research Committee of Eurocrine^®^ and the ESES and the Board of Eurocrine^®^.

The methods used to assess and improve overall data quality are as follows (Figure 1):

1. Real-time checks Data entries in Eurocrine are primarily based on drop-down lists with predefined categories. Free-text inputs are avoided whenever possible. Nevertheless, numeric values must be entered manually, which may result in errors. In addition, several contradiction checks are performed, and some variables are shown only if specific values of other variables are entered to prevent nonlogical value combinations. Owing to the high number of variables (>400), exhaustive plausibility checks cannot be embedded in the GUI of Eurocrine^®^. Furthermore, some value combinations may represent clinically unlikely, although not impossible, scenarios. These require global-level database analysis beyond the scope of GUI-based checks.
2. Understanding of variables Questionnaires are sent to the “doctors in charge” (a named responsible person for Eurocrine^®^ in a clinic) are used to clarify how they handle data entry in certain ambiguous situations.
3. Follow-up and outcome management Reminder emails are sent to “doctors in charge” if mandatory fields are missing or if the outcome of postoperative complications remains unclassified. For example, the complication ‘vocal cord paresis’ requires a complex evaluation across multiple variables and time points to distinguish between temporary and permanent outcomes. Complex algorithms account for pre-and postoperative assessments of vocal cord function at different time points as well as the intraoperative use of neuromonitoring to classify all possible value combinations in a limited number of outcomes. The resulting outcome value is used to prompt reminder emails to the user if a complication was entered but not evaluated during follow-up visits.
4. Interactive reports for internal audits Interactive data quality reports were developed in Microsoft Power BI for decentralized internal audits at the clinic level. These allow users/clinics to access and assess their data at any time and place through the Microsoft (MS) Power BI service. The interactive reports allow filtering of all variables and patient cases, making it easier to identify missing data, contradictory data, range violations, outliers, and implausible data. The identified records can then be crosschecked by the “doctor in charge” to review the patient chart and corrected via the Eurocrine^®^ web interface. Help texts guide the use and interpretation of the reports. In addition, predefined standard reports and benchmark summarize clinic performance with visual graphs, providing real-time feedback for quality improvement.
5. External audits External data audits began in Switzerland in 2019 on the basis of a national agreement. Between 2022 and 2025, the positive experience in Switzerland led to an extension of audits to Türkiye, Greece, Germany, and Spain. Following a revision of Eurocrine’s statutes in 2019, all participating clinics agreed to be audited at random intervals. Since then, three clinics per year were randomly chosen to be audited by the Vienna Eurocrine^®^ office. During each audit, 10 variables from 20 patients are cross-checked against clinic records. The selection of patients and procedures is conducted randomly, and the results are blinded to both the auditors and the clinics to preserve objectivity. In Sweden, regular audits in 3 to 5 clinics per year have been taking place since the commencement of the registry.
6. Encompassing data quality assessment Eurocrine data management complements the previous approaches through centralized data quality monitoring via the R package dataquieR [2, 10]. This tool generates standardized HTML reports across a broad series of indicators, e.g., completeness (e.g., missing values), consistency (e.g., range violations, contradictions), and accuracy (e.g., univariate outliers, multivariate outliers, time trends, centre effects). DataquieR reports are created in two steps: first, all the requested data quality aspects are computed; second, all the results are compiled into a structured HTML output.

**Fig. 1.**
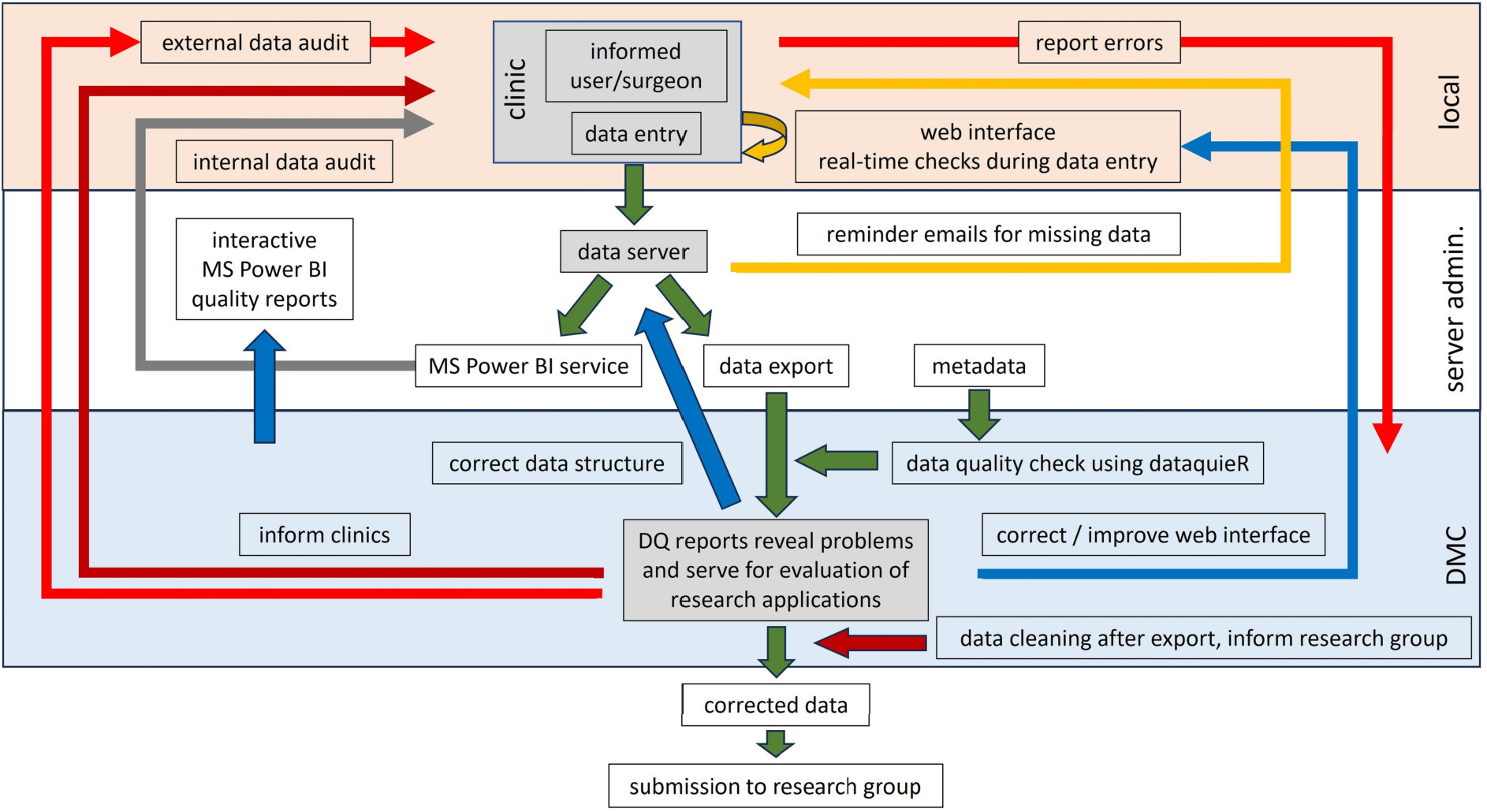
Workflow of data quality measures in the Eurocrine registry. Figure legend: Data quality assurance in Eurocrine® is implemented on the following different levels. First, the web interface used for data entry includes real-time inconsistency checks and limits settings to prevent erroneous input. If mandatory data or follow-up data are missing, the member clinic is notified by e-mail. Second, data are transferred to the MS Power BI service every night, allowing access to predefined interactive quality reports and standard reports with clinic-specific data at any time. These support the clinic’s internal data audits facilitated by MS Power BI reports designed by the Data Management Committee (DMC). Third, routine data exports and analyses with dataquieR are used to assess overall data quality within the DMC. The results from MS Power BI and dataquieR reports are used to correct the data and improve the web interface. Member hospitals are informed when local data quality issues are detected. In addition, external data audits verify the accuracy of the selected data through random sampling. DataquieR reports are also used to inform research groups receiving data exports in accordance with approved research projects.

## Results

The methods described above revealed various data quality issues, leading to multiple database software revisions, user notifications via newsletters, and both decentralized and centralized data corrections. Exemplary results of these data quality analyses are presented below.

### Questionnaires

In 2021, a questionnaire was distributed to participating clinics to evaluate how data entry was performed. One of the questions addressed postoperative calcium or vitamin D medication: “When do you enter ‘Calcium and Vitamin D-Medication for parathyroid insufficiency’ following thyroid and parathyroid surgery?”. Although the phrase “for parathyroid insufficiency” was intended to indicate that the variables specifically referred to the clinical situation of postoperative hypoparathyroidism, the answers revealed inconsistent interpretations. Some clinics routinely use calcium or vitamin D supplementation, since patients are discharged early on day 1 or 2 post surgery, and the need for treatment of hypoparathyroidism may not be obvious at this time point. As a result, new sub-variables were added to clarify the reasons for the medication (Figure 2).

**Fig. 2.**
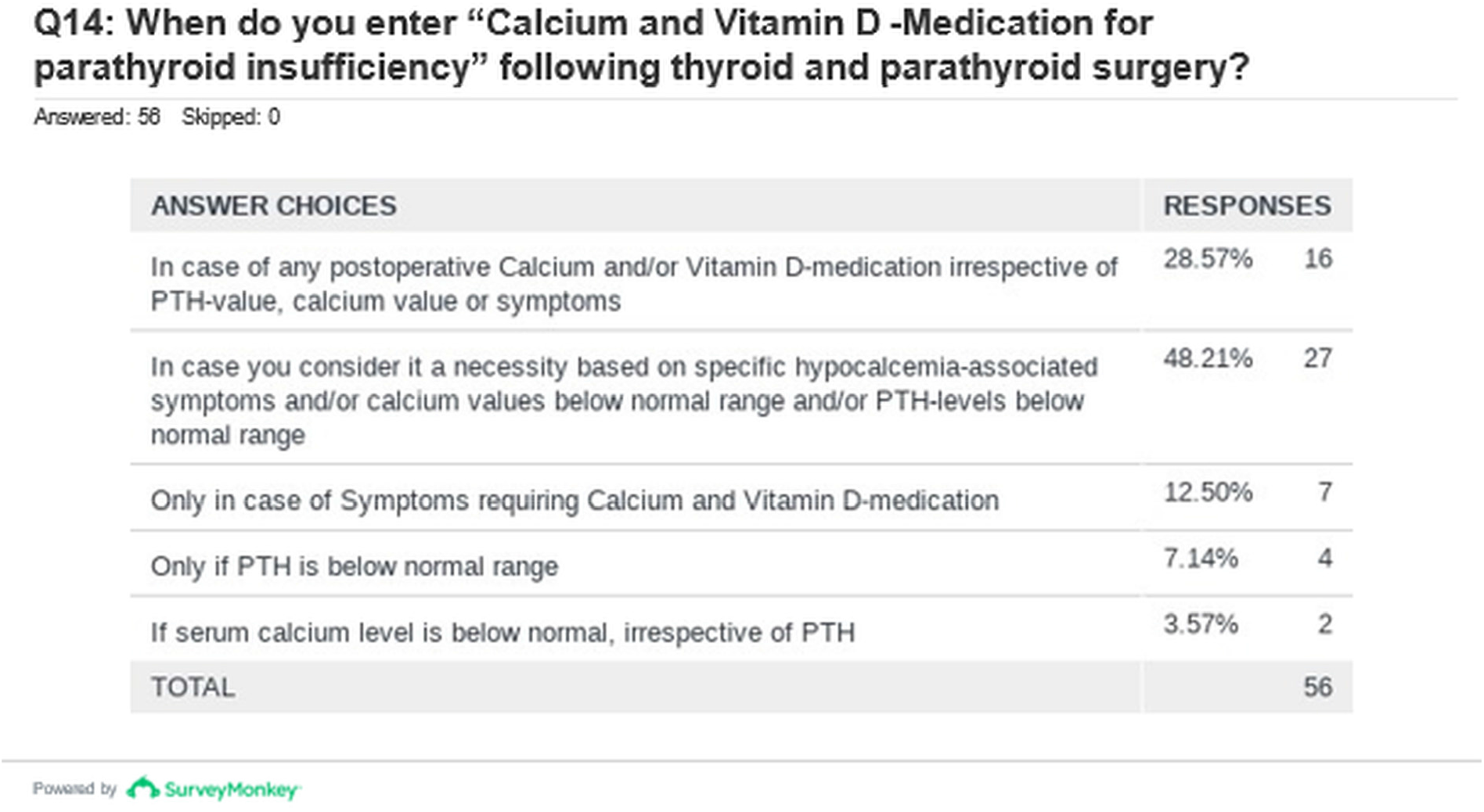
Results for an example item from the questionnaire on data entry assessment. Figure legend: The results for an example item from the questionnaire on data entry assessment highlight the varied interpretation of the mandatory variable “calcium or vitamin D medication for parathyroid insufficiency”. The responses revealed that the “yes” or “no” inputs were inconsistent due to the unclear definition of hypoparathyroidism. As a result, additional variables were added to clarify the clinical rationale for the use of calcium or vitamin D medication.

### MS Power BI quality reports and dataquieR reports

After one year of data collection, reviews of the global database using MS Power BI reports revealed incorrect data for dates and serum calcium values (Figure 3). To address this and prevent implausible data entry, real-time plausibility checks were implemented at the web interface using limits for all numeric values and dates. Similar errors were discovered for the numerical value ‘number of tumors’, which refers to the number of tumors in the thyroid. Typing errors and misinterpretation of the variable’s meaning led to implausible data.

**Fig. 3.**
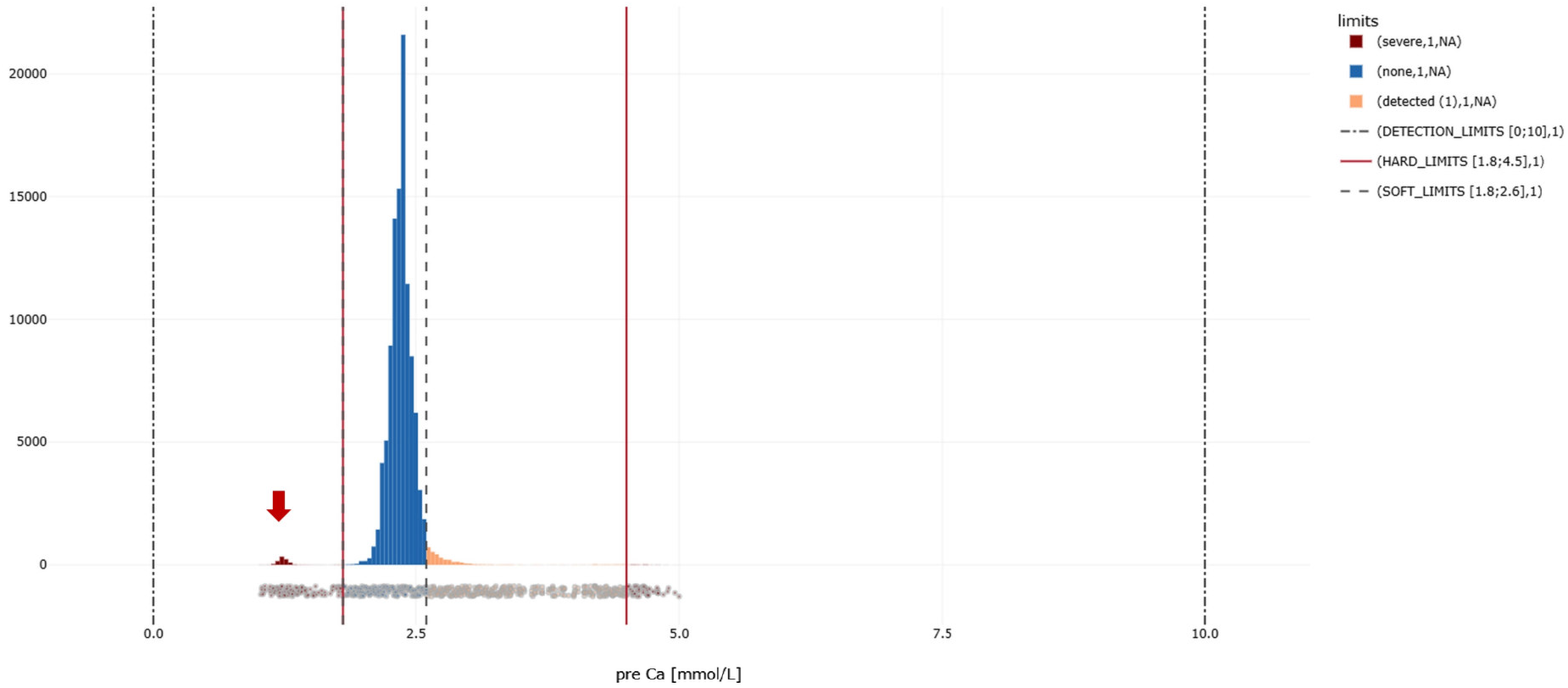
Output for limit deviations for preoperative serum calcium levels [mmol/l] using dataquieR. Figure legend: Serum calcium levels are entered as numeric values into the Eurocrine^®^ web interface. Input is possible via two scales: total calcium or ionized calcium in mmol/l or mg/dl. After a value is entered in any of the four fields, the 3 other entries are calculated automatically. However, if the unit or total calcium and ionized calcium are mixed, this results in incorrect values falling outside physiologically expected ranges. Histograms are helpful for detecting such cases. The frequency distribution of the values for preoperative serum calcium is shown in the graph. The vertical red lines represent *a priori* defined physiological limits (hard limits=inadmissible values), whereas the vertical dashed lines represent the limits of normal values (soft limits=uncertain values). The bars marked with red arrows below the lower (left) hard limit likely represent values that can be attributed to an incorrect entry. Entries outside the defined hard limits have no longer been possible since 2017, suggesting that the detected errors occurred earlier.

Additionally, inadmissible values and implausible data were identified via dataquieR. Inadmissible values, which should have been excluded by data entry checks in the web interface, were still found. This was related to programming errors and concerned variables with dependencies (i.e., main variables and subordinate variables, applicable only under specific main variable categories). In addition, the change in the TNM classification from the 7th to the 8th editions led, albeit in very rare cases, to incorrect assignments of T stage categories. These errors were corrected and have been completely avoided thereafter.

### Decentralized auditing

Since its introduction in 2019, three clinics per year have been randomly chosen to be audited by the Vienna Eurocrine^®^ office. The audits are conducted by representatives from other Eurocrine^®^ clinics familiar with the database. Thus, they are well aware of potential difficulties in interpreting variable nomenclature. Since the introduction of audits, nine clinics have been audited. The average percentage of correct items was 95.9% (range: 89% - 99%). Fifteen of the 17 clinics were awarded a Eurocrine^®^ certificate for data validity. These certificates are issued by the Eurocrine^®^ Society if more than 90% of the checked items are correct. Data checks in Sweden also showed an average accuracy and completeness rate of 94%. Ninety-five percent of all thyroid procedures are documented in Eurocrine^®^.

### Centralized data quality evaluation

The centralized assessment of data quality using dataquieR revealed no issues related to data integrity, and all mandatory data elements were almost 100% complete (Figure 4). In the same way the completeness of the segments basic data, preoperative data, operative data, postoperative data and the first follow-up revealed only 0.86% and 1.8% missing data for the segments “postoperative” and “first follow-up”. Since data export represents a snapshot of ongoing data entry, and the in-hospital care of some patients may not have been completed at the time of the data export, an overall 100% completeness cannot be achieved.

**Fig. 4.**
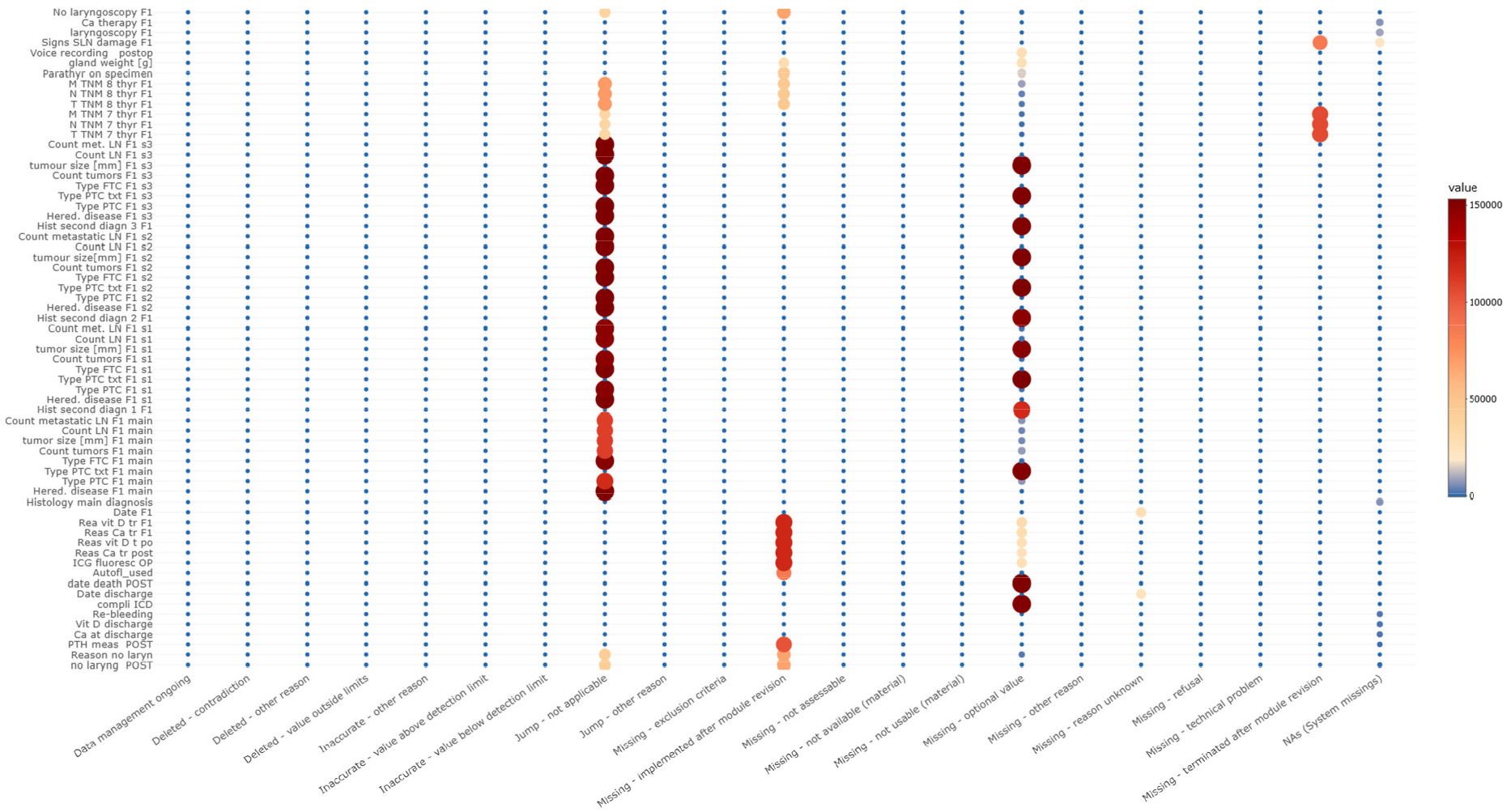
Overview of missing data via dataquieR. Figure legend: Heatmap for missing values of selected registry variables. The percentage of missing values is represented by the size and color of the dots, as shown in the legend. dataquieR displays annotated causes of missing values, which are defined in a code table where each cause is assigned a unique value. If a value is missing or falls outside an expected range (e.g., negative numbers for a body height), the value is replaced with a corresponding value of the code table. For example, in the case of benign thyroid diseases, the TNM classification is not applicable, and values must be missing. These scenarios are summarized in the column ‘Jump – not applicable’. Other reasons include the removal of variables or the introduction of new variables, such as the transition from the 7^th^ to the 8^th^ editions of the TNM system. These cases are depicted in the columns ‘Missing – implemented after module revision’ and ‘Missing – terminated after module revision’. Optional data entries are displayed in the column ‘Missing – optional value’. Missing values that are not referenced in the code table are shown in the column “NAs (System missings)”, indicating the presence of unclassified missing values.

Checks for inadmissible values revealed limit deviations in less than 1.3% of the numeric data elements and dates. These errors occurred mainly during the early phase of data entry, when limit checks had not yet been implemented in the web interface. For some data elements, inadmissible categorical values were caused by programming errors, which were resolved along with the corresponding data corrections.

Encompassing analysis of contradictions demonstrated only minor logical deviations, i.e., less than 0.6% (Figure 5). dataquieR analyses revealed no relevant findings for data accuracy.

**Fig. 5.**
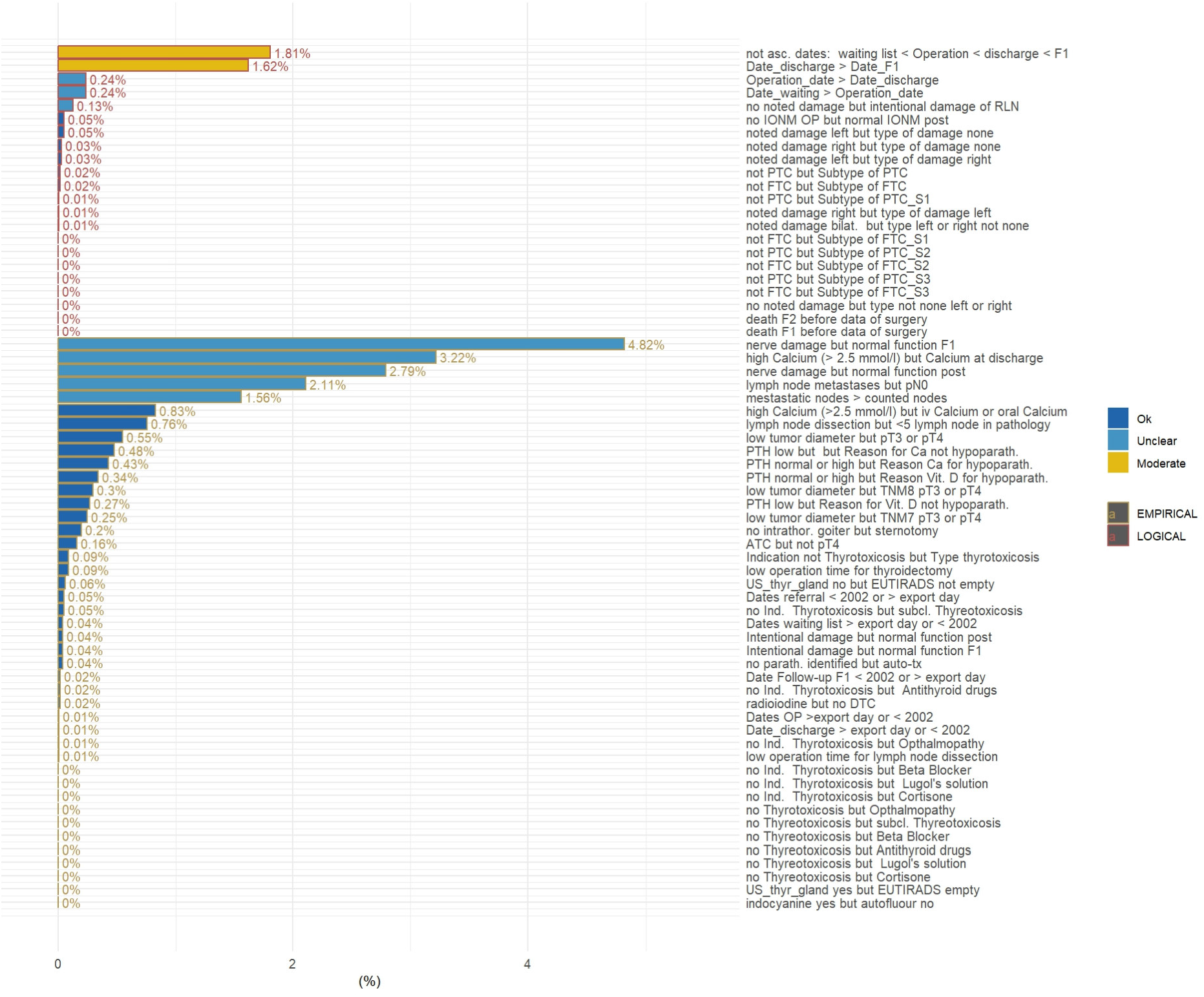
Evaluation of logical and empirical contradictions via dataquieR. Figure legend: The figure shows an assessment of empirical and logical contradictions using 65 different rules, defined according to REDCAP syntax. The higher the number or percentage of rule violations is, the lower the data quality. For the classification of observed logical and empirical contradictions into up to 5 classes (ok, unclear, moderate, important, critical) grading rulesets are used that can be customized. All but two types of logical contradictions are below 1%, which indicates a sufficient level of data quality. The two most frequently occurring logical contradictions “not asc. dates: waiting list < Operation < discharge < F1” and “Date_discharge > Date_F1” were due to missing real-time checks of the web interface in the early phase of Eurocrine^®^. Other logical contradictions involved entries of dependent variables. Such data entry errors were possible before the implementation of real-time validation. Following the detection of these logical contradictions, the web interface was changed. Empirical contradictions are related to unlikely but theoretical possible scenarios. For such contradictions, rule violations with a frequency of more than 1% may still be acceptable. The observed frequencies below a threshold of 5% are within the expected range.

## Discussion

Medical registries have the potential to provide profound insights into clinical treatment quality, i.e., so-called ‘real-world’ outcome data. For this reason, registry data serve as an important source of information not only for the development of guidelines and treatment recommendations but also for health ministries, health insurance companies, and medical companies.

The design and implementation of a new medical registry is extremely complex. This is especially true when numerous data elements are needed to capture a multitude of clinical scenarios associated with different treatment routines across clinics and countries. Despite meticulous planning and testing, the production phase will likely reveal problems that were not foreseen during registry development. Changing clinical routines and new scientific questions, as well as evolving classifications (such as the TNM system or the WHO definition of diseases), challenge the relevance of the collected data and necessitate adaptations of the database. While this requires the registry to be flexible, especially when data are collected over a long period, implemented changes may compromise data consistency and introduce systematic errors.

Routine, ongoing data quality evaluation is therefore indispensable for medical registries. Ensuring high data quality starts with informing and training users. Prompt input of the data at the time of admission, operation, and discharge of a patient, supported by clear, easy-to-understand input masks with integrated plausibility queries, is optimal. Despite these efforts, errors and unintentional and potentially intentional false data entries can occur. Feedback to the user regarding data issues, as provided in Eurocrine^®^, through MS Power BI quality reports, which allows users the possibility of correcting errors. However, it is not sufficient to provide only a tool for quality checking: it must also be actively used at both the clinical and national levels to positively impact data quality. The motivation and dedication of surgeons in participating clinics is an essential prerequisite for this. The motivation depends on the benefits the registry offers to individual clinics and surgeons. Examples include the possibility of evaluating clinic-specific outcome quality, benchmarking, and participation in research studies.

Furthermore, we describe here for the first time the value of the R package dataquieR in generating comprehensive data quality reports for a clinical registry. The results of these assessments support registry administrators in identifying problems that require further action. The dataquieR reports for the Eurocrine^®^ thyroid module revealed very good data quality regarding completeness, consistency, and accuracy, supporting the efficiency of previous data quality assurance measures. Future reports that encompass the complete database of Eurocrine^®^ will support the joint Research Committee of Eurocrine^®^ and ESES when evaluating applications for data extraction for retrospective studies and prospective trials. Clinic-specific reports using dataquieR and MS Power BI are equally suitable as a basis for onsite data audits and in the accreditation processes for endocrine surgery centers by the European Society of Endocrine Surgeons (ESES). This process is currently under development. Several publications based on Eurocrine® data underlined the relevance of the data for contemporary scientific issues in the field of endocrine surgery [11–20].

The impact of decentralized use of the standard reports, benchmark reports, and data quality reports in the MS Power BI service cannot be analysed in detail since the connection between the Eurocrine® database and the MS Power BI service is unidirectional. Data cannot be changed in the MS Power BI service for security reasons. Only the identification of data issues is supported. Changes or corrections of data by a clinic can be performed at any time via the Eurocrine® Web-Interface. The reason for a change cannot be entered until now, and snapshots of the database are not routinely stored. However, usage metrics of the MS Power BI service as well as results from clinic audits indicated that surgeons in participating Eurocrine^®^ clinics are highly committed to achieving and maintaining excellent data quality.

DataquieR and MS Power BI quality reports have limitations. MS Power BI quality reports are used mainly by clinics through cloud-based MS Power BI service since usage by local computers requires the installation of additional software. MS Power BI allows the customization of visuals and the design of custom visuals that contain coding in R or Python. While this opens the door to unlimited possibilities of data visualization, it also has drawbacks. Small volumes (several thousand observations) of data can be managed with predefined custom visuals, but owing to the storage requirements of tools such as “plotly,” the service capacity is often exceeded with large volumes of data (> 10,000 observations).

The first versions of dataquieR did not meet all the requirements for analysing data quality in a complex medical registry. However, through close cooperation with developers, adjustments and updates could be made, allowing for optimized and comprehensive reports [10, 21]. An essential prerequisite for the use of dataquieR is a detailed description of the metadata. To enable the necessary [21] wide scope of assessments, it is mandatory to define data types, expected distributions, value ranges, expected categorial values, missing data labels, contradiction rules, and other information. Creating metadata in XLSX sheets requires precise adherence to the formatting requirements specified by dataquieR, which are documented in detail. In the most recent version, it is possible to adjust the quality grading for each variable and data quality dimension. For large databases, such as Eurocrine^®^, report generation requires substantial hardware and computing time: generating a report for 150,000 observations and 190 variables requires more than 5 hours of computation, despite parallelization, and more than 6 hours for report rendering on a PC with 128 GB of core memory and a Core i7 processor with 12 cores.

It is important to note that data quality evaluation in medical registries must be tailored to the specific needs and requirements of each registry. There is no one-size-fits-all solution, and the choice of methods and tools should be carefully considered.

We believe that the procedures implemented in Eurocrine^®^ can serve as both a guide and a template for other registries. Building on foundations such as those presented here could ultimately contribute to the development of a widely accepted standard for assessing data quality in medical registries. Such a standard is urgently needed, as medical publications generally provide little or no information on data collection methods, data quality analysis, or data cleaning. Nevertheless, these studies are frequently included in systematic reviews without critical appraisal of their underlying data sources or quality. This omission is problematic, as published findings based on registry data can strongly influence the development of clinical guidelines. To address this, structured comprehensive data quality reports should be included in the review process of any medical publication on the basis of registry data.

## Data Availability

The data that support the findings of this study are not openly available due to reasons of sensitivity and ongoing studies. The data are available from the Eurocrine Board upon reasonable request. Region Skane, Sweden (Scania County Council), is the organisation responsible for processing data. Further information is provided at https://eurocrine.eu/.

## Declarations

### Ethics approval and consent to participate

EUROCRINE is subject to the General Data Protection Regulation (GDPR) including national implementation of the legislation. Consent to participate in the Eurocrine® Registry was obtained according to National Regulations and Laws for all patients. Further, all patients were informed of their rights according to GDPR. Informed consent was obtained from all patients in Countries in which the local Institutional Review Board (IRB) demanded such informed consent.

All procedures performed were in accordance with the ethical standards of the institutional and national research committee and with the 1964 Helsinki declaration and its later amendments or comparable ethical standards. In Germany the registry was approved by the Ethics Committee of the Rhineland-Palatinate Medical Association (processing number: 837.119.15 (9887)).

The registry is listed in the German Clinical Trials Register (DRKS00009246, Registration Date September 1^st^ 2015, https://drks.de/search/en/trial/DRKS00009246).

## Consent for publication

All authors agree with the publication of the manuscript.

## Availability of data and materials

The data that support the findings of this study are not openly available due to reasons of sensitivity and ongoing studies. The data are available from the Eurocrine Board upon reasonable request. Region Skåne, Sweden (Scania County Council), is the organisation responsible for processing data. Further information is provided at https://eurocrine.eu/.

## Authors’ contributions

T.M., A. B., and T. C. are members of the Eurocrine Board and the Data Management Committee of Eurocrine responsible. In this function they developed measures to assure the data quality in the registry. Moreover, they drafted the manuscript. S. S. and C. O. S. developed the R package dataquieR and advised its application. Both reviewed and revised the manuscript.

